# Sleep Disorders Modify the Age-Related Trajectory of Circadian Rest-Activity Rhythms: Evidence from NHANES 2011–2012 Wrist Actigraphy

**DOI:** 10.64898/2026.05.28.26354369

**Authors:** Luke Yin, Cooper Lee, Andrew Wong

## Abstract

**Background:** Circadian rest-activity rhythms weaken with age, but whether sleep disorders modify this trajectory is unknown.

**Methods:** We analyzed wrist accelerometry data from 4,386 participants aged 6–80 years in the 2011– 2012 National Health and Nutrition Examination Survey (NHANES). Circadian features were extracted using cosinor analysis and nonparametric methods; a Circadian Disruption Index (CDI) was constructed from five standardized components. Survey-weighted regression with natural cubic splines and Wald F-tests tested age-by-sleep-disorder interactions using Taylor series linearization for variance estimation.

**Results:** Doctor-diagnosed sleep disorder (*N* = 360, 8.2%) was associated with significantly different age-related trajectories of amplitude (*F*(2, 17) = 11.24, *p* = 0.0008) and MESOR (*F*(2, 17) = 8.22, *p* = 0.0032), both surviving Bonferroni correction (*p* < 0.006). CDI was higher in those with a sleep disorder (0.290 vs. 0.131, *p* < 0.001) and was independently associated with higher BMI (*β* = 1.33 kg/m^2^, *p* < 0.001), higher HbA1c (*β* = 0.089%, *p* = 0.004), greater diabetes prevalence (*β* = 3.8 percentage points, *p* < 0.001), and worse depressive symptoms (*β* = 0.43 PHQ-9 points, *p* = 0.020). Sensitivity analyses using a broader sleep problem exposure did not replicate these interactions.

**Conclusions:** Doctor-diagnosed sleep disorders are associated with an altered age-related decline in circadian amplitude and mean activity level. CDI was independently linked to cardiometabolic and depressive outcomes, supporting a mechanistic connection between clinically significant sleep pathology and circadian disruption across the lifespan.

## 1 Introduction

Circadian rest-activity rhythms, reflecting the approximately 24-hour oscillation between periods of activity and rest, are a fundamental feature of human physiology [1]. Their strength and regularity are maintained by the central circadian pacemaker in the suprachiasmatic nucleus (SCN) and entrained by photic and non-photic cues including light, social schedules, and feeding patterns [2–5]. A substantial body of evidence demonstrates that these rhythms weaken progressively with age: older adults exhibit lower amplitude, reduced interdaily stability, increased fragmentation, and earlier acrophase compared to younger individuals [6–9]. This age-related circadian attenuation is partly attributable to neuronal desynchrony within the aging SCN and diminished responsiveness to photic entrainment cues [10, 11]. The downstream consequences include impaired cognitive function, increased cardiometabolic risk, and greater all-cause mortality [12–15].

Sleep disorders affect an estimated 50 to 70 million Americans [16] and encompass a heterogeneous group of conditions including insomnia disorder, obstructive sleep apnea, restless legs syndrome, and circadian rhythm sleep-wake disorders [17–20]. These conditions share mechanistic features that could plausibly disrupt circadian organization. Recurrent nocturnal hypoxemia in sleep apnea activates inflammatory and autonomic pathways that interfere with peripheral circadian clock gene expression [21–23]. Chronic insomnia is associated with heightened cortical and autonomic arousal that fragments rest-activity rhythms even when total sleep duration is preserved [24, 25]. Circadian rhythm disorders by definition represent direct pathology of the timing system itself [26, 27]. Despite these mechanistic connections, few large-scale population studies have examined whether a clinical sleep disorder modifies the normal agerelated trajectory of objectively measured rest-activity rhythms.

Wrist actigraphy provides an accessible, objective measure of rest-activity rhythms at population scale [28– 30]. The 2011–2012 National Health and Nutrition Examination Survey (NHANES) included a wrist actigraphy component in a nationally representative sample, enabling population-level estimation of circadian features across a wide age range [31, 32]. Standard cosinor modeling yields amplitude, acrophase, and MESOR as indices of rhythm strength, timing, and mean level [33, 34], while nonparametric approaches provide complementary characterization of fragmentation and stability [35, 36]. Composite disruption indices derived from multiple circadian metrics have been shown to associate with cognitive decline, depression, and metabolic disease [12, 13, 37, 38]. The finding that circadian misalignment per se, independent of reduced sleep duration, adversely affects glucose metabolism and cardiovascular biomarkers further motivates population-level study of circadian disruption [39–42].

The primary aim of this study was to test whether doctor-diagnosed sleep disorder modifies the age-related trajectory of rest-activity circadian metrics in a nationally representative US sample, using survey-weighted regression with Taylor series linearization to properly account for the NHANES complex sampling design [43, 44]. The secondary aim was to examine associations between a CDI and cardiometabolic and mental health outcomes to validate the clinical relevance of the composite. We hypothesized that clinically diagnosed sleep disorders would be associated with a different pattern of agerelated circadian attenuation, given the shared biological substrates of circadian organization and sleep pathology [45–47].

## 2 Materials and Methods

### 2.1 Study Design and Population

We used data from the 2011–2012 NHANES cycle, a cross-sectional, nationally representative survey of the non-institutionalized US civilian population conducted by the National Center for Health Statistics [31]. NHANES uses a complex, multistage probability sampling design with oversampling of certain demographic subgroups. All participants provided written informed consent, and the protocol was approved by the NCHS Research Ethics Review Board.

Of 9,756 participants who completed the mobile examination center visit, 6,917 were fitted with a wristworn accelerometer (ActiGraph GT3X+) and asked to wear it for seven consecutive days. After excluding participants with fewer than four valid wear days (defined as at least 600 wear minutes per day with quality flag = 0), 5,610 participants remained. Of these, 4,386 had complete 24-hour activity profiles and non-missing values for the primary exposure and all model covariates, constituting the analytic sample. The sample included individuals aged 6 to 80 years, encompassing childhood through older adulthood; this broad age range was retained to maximize statistical power for the age-by-sleep-disorder interaction and to characterize circadian trajectories across the full lifespan. The sleep disorder exposure (SLQ060) is administered to NHANES participants aged 16 and older; participants aged 6–15 were classified as not having a doctor-diagnosed sleep disorder, as this question was not applicable to them. Cardiometabolic outcome models (BMI, HbA1c, diabetes, PHQ-9) were adjusted for age as a continuous covariate to account for developmental differences across the age range.

### 2.2 Actigraphy Processing

Minute-level accelerometry data were read from the NHANES PAXMIN G file [32]. Valid wear minutes were identified using the device quality flag (PAXQFM = 0). Clock time for each sample was computed by adding the device-recorded day start time (PAXMSTD, minutes since midnight) to the within-day sample offset in minutes, calculated as (PAXSSNMP − cumulative day start sample)*/*4800, where 4800 reflects the 80 Hz sampling rate multiplied by 60 s. Activity was measured as vector magnitude in milli-g units (PAXMTSM). Hourly mean activity was computed for each valid day and then averaged across valid days to produce a single 24-hour profile. Only participants with complete profiles across all 24 h were retained. This approach follows established NHANES actigraphy processing conventions [30, 32].

### 2.3 Circadian Feature Extraction

#### Cosinor analysis

A single-component cosinor model was fitted to each participant’s 24-hour profile: *y*(*h*) = MESOR + *A* cos(2*πh/*24) + *B* sin(2*πh/*24), estimated by ordinary least squares [33, 34]. This yields the MESOR (midline-estimating statistic of rhythm; mean activity level), amplitude (half the peak-to-trough range, 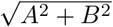), acrophase (clock time of predicted peak activity, arctan 2(−*B, A*) × 24*/*(2*π*) mod 24), and goodness-of-fit (*R*^2^).

#### Nonparametric metrics

Interdaily stability (IS) quantifies day-to-day regularity of the rest-activity pattern [9, 35]. Intradaily variability (IV) quantifies within-day fragmentation of the rhythm. The 10-h most active window (M10) and 5-h least active window (L5) capture the timing and intensity of peak active and rest periods, respectively. Relative amplitude (RA = (*M* 10 − *L*5)*/*(*M* 10 + *L*5)) summarizes contrast between active and rest periods [9]. Higher IS and RA and lower IV indicate more robust rhythmicity.

#### Circadian Disruption Index

The CDI was constructed as the unweighted mean of five standardized (z-scored) components, each oriented so that higher values indicate greater disruption: −*z*(amplitude), −*z*(cosinor *R*^2^), *z*(IV), −*z*(IS), and −*z*(RA). This composite approach summarizes multiple dimensions of circadian organization into a single scalar index suitable for association analyses [12, 37].

### 2.4 Sleep Disorder Classification

The primary exposure was doctor-diagnosed sleep disorder, defined as an affirmative response to “Have you ever been told by a doctor or other health professional that you have a sleep disorder?” (SLQ060). This variable captures clinically recognized sleep pathology encompassing insomnia disorder, obstructive sleep apnea, restless legs syndrome, and circadian rhythm sleep disorders as classified by the International Classification of Sleep Disorders [27]. For sensitivity analysis, a composite exposure (“any sleep problem”) was defined as an affirmative response to SLQ060 or “Have you ever told a doctor or other health professional that you have trouble sleeping?” (SLQ050).

### 2.5 Covariates and Outcomes

Covariates included age (years, continuous), sex, race/ethnicity (Non-Hispanic White, Non-Hispanic Black, Non-Hispanic Asian, Mexican American, Other Hispanic, Other), body mass index (BMI, kg/m^2^), and self-reported usual weekday sleep duration in hours. These covariates were included in all age-by-sleep-disorder interaction models. Secondary outcomes examined for association with CDI included BMI, glycated hemoglobin (HbA1c, %), diabetes diagnosis (DIQ010), and depressive symptoms (PHQ-9 score, continuous); CDI outcome models were adjusted for age, sex, and BMI.

### 2.6 Statistical Analysis

Survey-weighted regression models were fitted with natural cubic splines for age (4 total knots, with 2 interior knots placed at quantiles of the age distribution) interacted with sleep disorder status. A natural cubic spline with 4 knots produces *K* − 2 = 2 free basis functions after applying natural boundary constraints, yielding a numerator *df* = 2 for the joint age-by-sleep-disorder interaction Wald test. Variance was estimated via Taylor series linearization, the standard method for NHANES complex survey analyses [43, 44], implemented as a custom sandwich estimator equivalent to the svyglm() function in R. Globally unique PSU identifiers were created by combining stratum and PSU codes. Denominator degrees of freedom were 17 (19 PSU clusters minus 2 strata in the analytic sample).

Bonferroni correction was applied for eight simultaneous Wald tests (one per circadian metric), yielding a corrected threshold of *p* < 0.006. All analyses were conducted in Python 3.14. Figures were produced with matplotlib.

## 3 Results

### 3.1 Sample Characteristics

The analytic sample comprised 4,386 participants (Table 1). The mean age was 46.8 years (SD 19.0), and 52.1% were female. Doctor-diagnosed sleep disorder was present in 360 participants (8.2%). Compared to participants without a sleep disorder, those with a diagnosis were older (53.5 vs. 46.2 years, *p* < 0.001), had substantially higher BMI (33.0 vs. 28.3 kg/m^2^, *p* < 0.001), and reported fewer hours of sleep per night (6.4 vs. 6.9 h, *p* < 0.001). Sex distribution did not differ significantly between groups (*p* = 0.366). Race/ethnicity differed significantly (*p* < 0.001), with Non-Hispanic White participants more prevalent in the sleep disorder group (45.8% vs. 35.3%) and Non-Hispanic Asian participants less prevalent (5.6% vs. 13.9%).

**Table 1:**
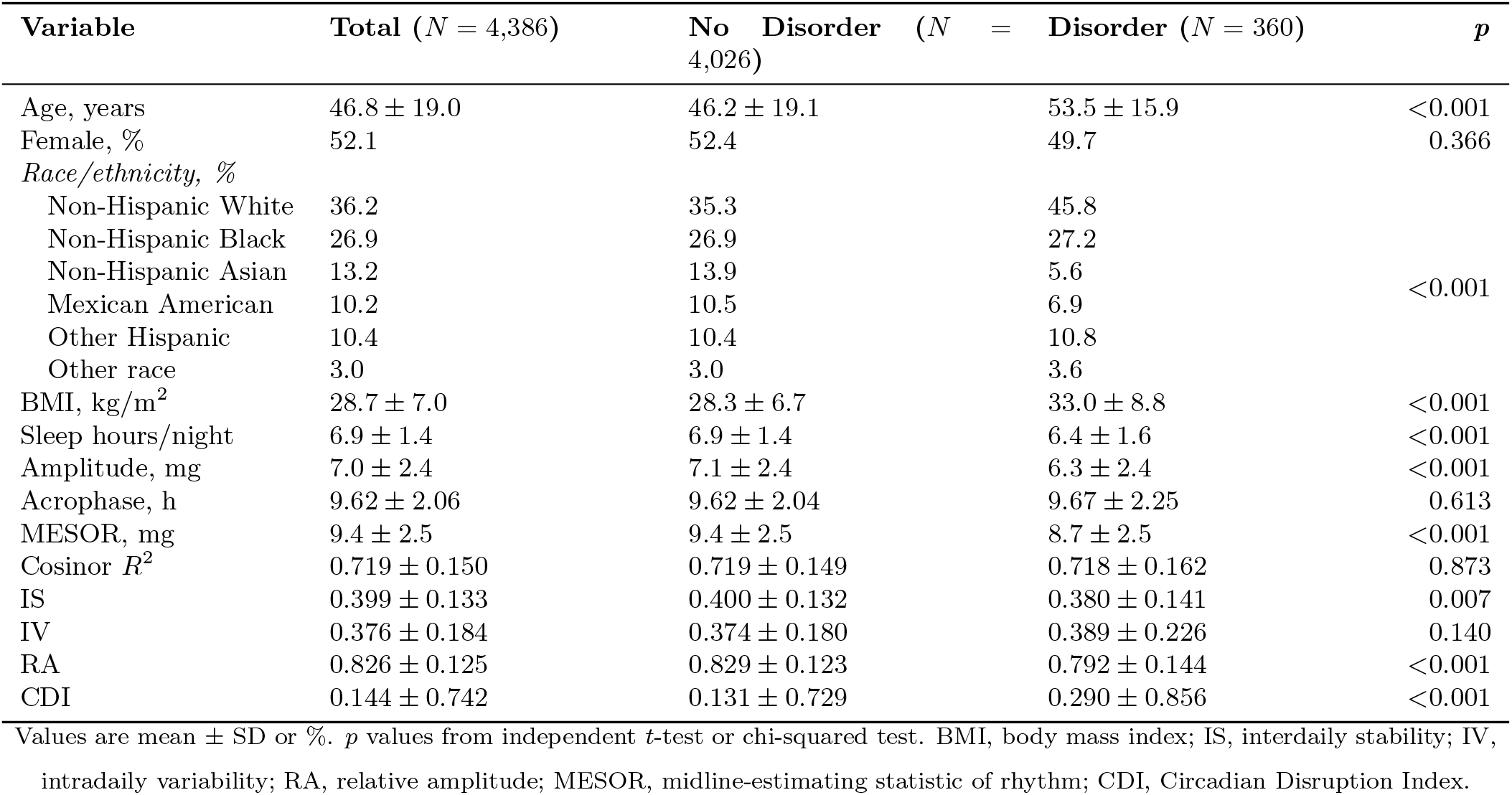
Participant characteristics by sleep disorder status.

### 3.2 Circadian Metrics by Sleep Disorder Status

Participants with a doctor-diagnosed sleep disorder showed lower activity amplitude (6.3 vs. 7.1 mg, *p* < 0.001), lower MESOR (8.7 vs. 9.4 mg, *p* < 0.001), lower interdaily stability (0.380 vs. 0.400, *p* = 0.007), and lower relative amplitude (0.792 vs. 0.829, *p* < 0.001) compared to those without a sleep disorder. Acrophase, cosinor *R*^2^, and IV did not differ significantly. CDI was substantially higher in the sleep disorder group (0.290 vs. 0.131, *p* < 0.001), indicating greater overall circadian disruption. Mean 24-hour activity profiles are shown in Figure 1.

**Figure 1:**
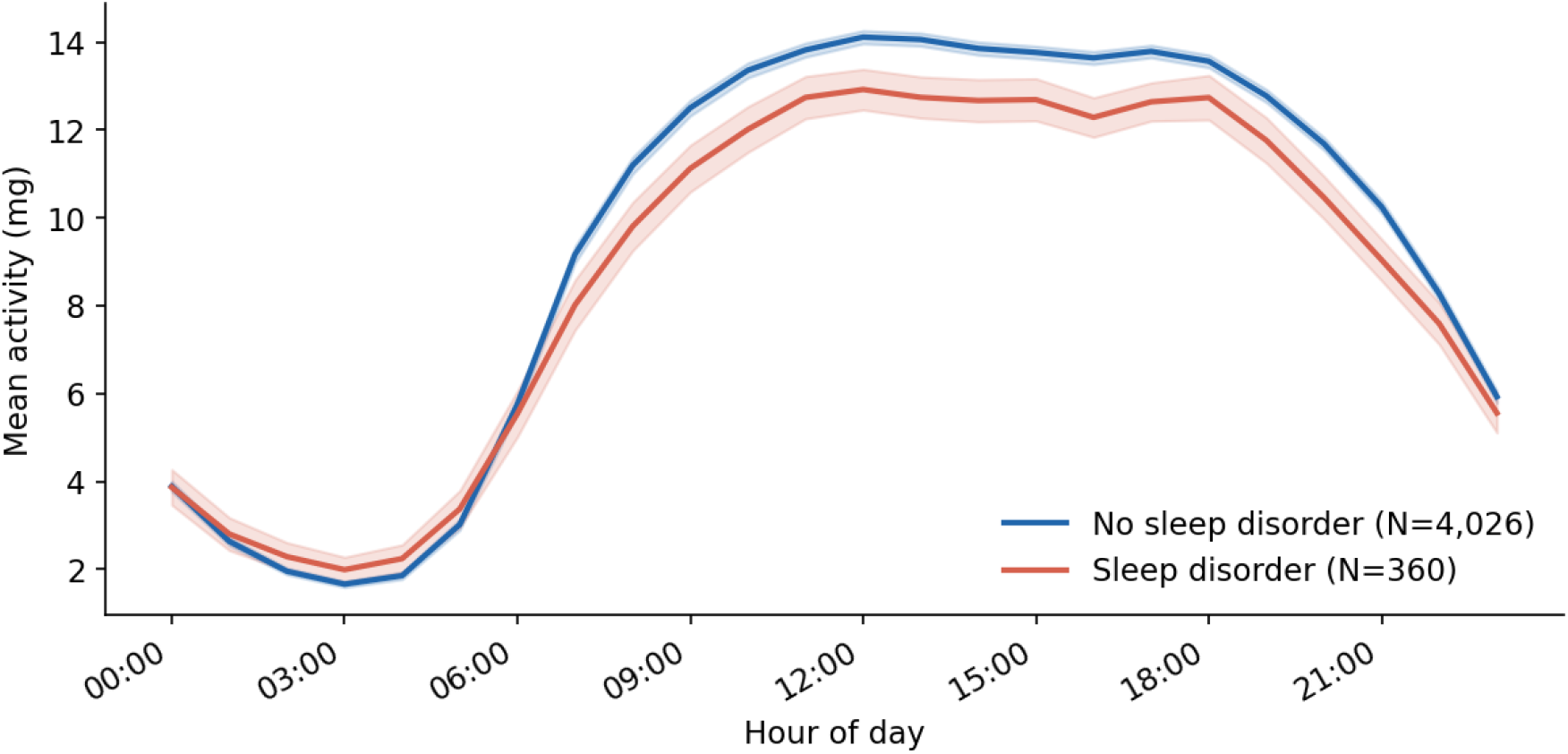
Mean 24-hour wrist accelerometry profiles by sleep disorder status. Blue line, no sleep disorder (*N* = 4,026); red line, doctor-diagnosed sleep disorder (*N* = 360). Shaded bands represent 95% confidence intervals. Activity is expressed as vector magnitude in mg.

### 3.3 Age by Sleep Disorder Interactions

Survey-corrected Wald test results for all eight circadian metrics are presented in Table 2. Two metrics showed significant age-by-sleep-disorder interactions after Bonferroni correction. Activity amplitude showed a strongly significant interaction (*F*(2, 17) = 11.24, *p* = 0.0008), and MESOR showed a similarly significant interaction (*F*(2, 17) = 8.22, *p* = 0.0032). Cosinor *R*^2^ showed a nominally significant interaction (*p* = 0.048) that did not survive Bonferroni correction. No significant interactions were observed for acrophase, IS, IV, RA, or CDI.

**Table 2:** Survey-corrected Wald F-tests for age × sleep disorder interactions on circadian metrics.

| Metric | F Statistic | df | p Value | Bonferroni Significant |
| --- | --- | --- | --- | --- |
| Amplitude | 11.24 | 2, 17 | 0.0008 | Yes *** |
| MESOR | 8.22 | 2, 17 | 0.0032 | Yes ** |
| Cosinor $R^2$ | 3.66 | 2, 17 | 0.048 | No * |
| Acrophase | 0.22 | 2, 17 | 0.802 | No |
| IS | 0.17 | 2, 17 | 0.844 | No |
| IV | 1.94 | 2, 17 | 0.174 | No |
| RA | 0.14 | 2, 17 | 0.870 | No |
| CDI | 2.28 | 2, 17 | 0.132 | No |

Survey-corrected Wald F-test for joint significance of the age × sleep disorder spline interaction. The natural cubic spline with 4 knots produces $K - 2 = 2$ free basis functions after natural boundary constraints, giving numerator $df = 2$ ; denominator $df = 17$ (19 PSU clusters minus 2 strata). Bonferroni threshold: $p < 0.006$ (8 tests). IS, interdaily stability; IV, intradaily variability; RA, relative amplitude; CDI, Circadian Disruption Index; MESOR, midline-estimating statistic of rhythm.

Predicted age trajectories with 95% confidence intervals are shown in Figure 2, illustrating that the sleep disorder group shows a different pattern of amplitude and mean activity level across the lifespan. A comparative summary of all Wald F-test results for the primary and sensitivity analyses is shown in Figure 3.

**Figure 2:**
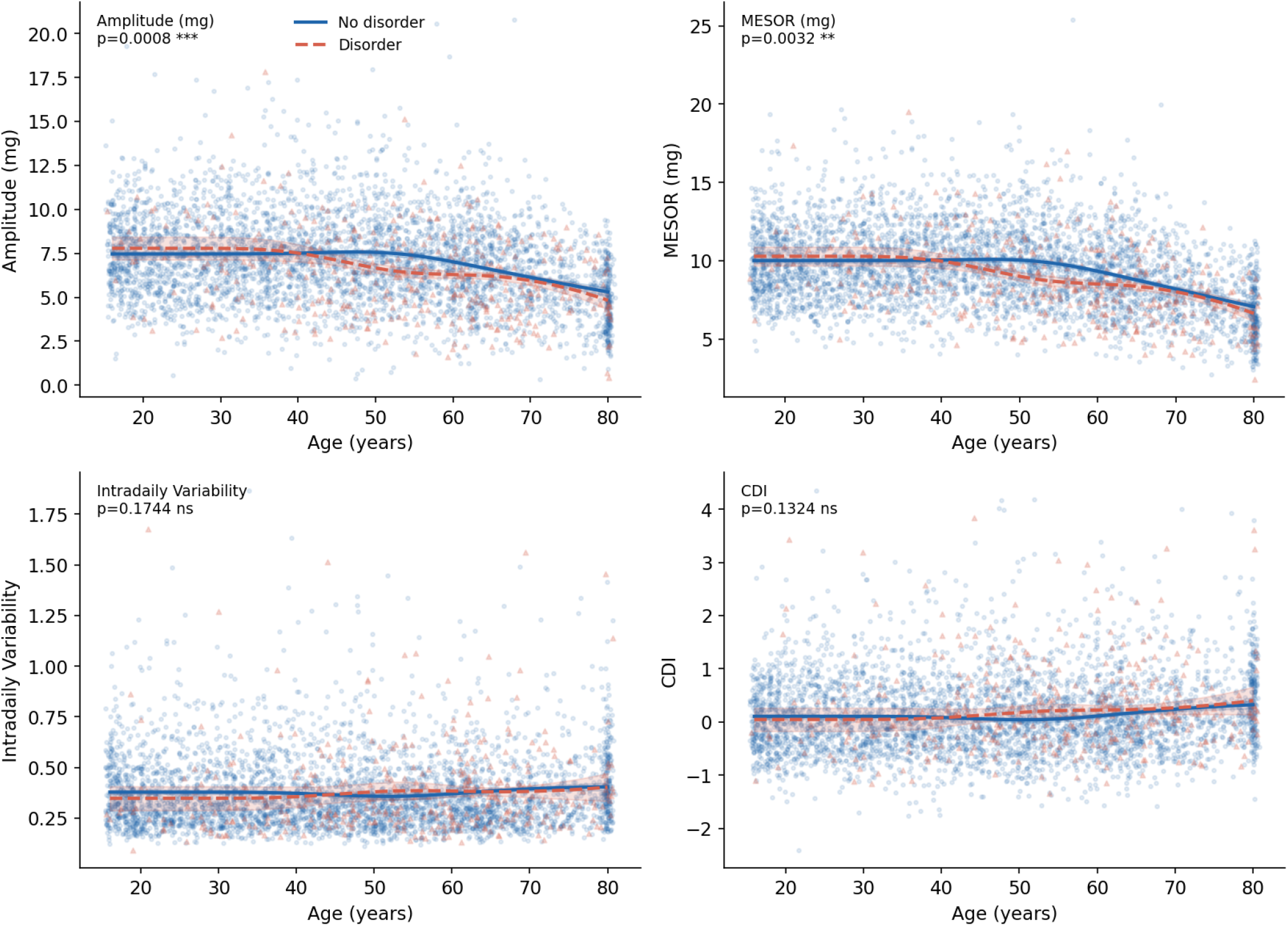
Age-related trajectories of key circadian metrics by sleep disorder status, with 95% confidence bands (shaded). Curves represent predicted values from survey-weighted regression with natural cubic spline terms for age (4 total knots, 2 interior) at mean covariate values. Points represent individual participant values (jittered). Survey Wald *p*-values for the joint age × sleep disorder interaction are shown. Blue, no sleep disorder; red, doctor-diagnosed sleep disorder.

**Figure 3:**
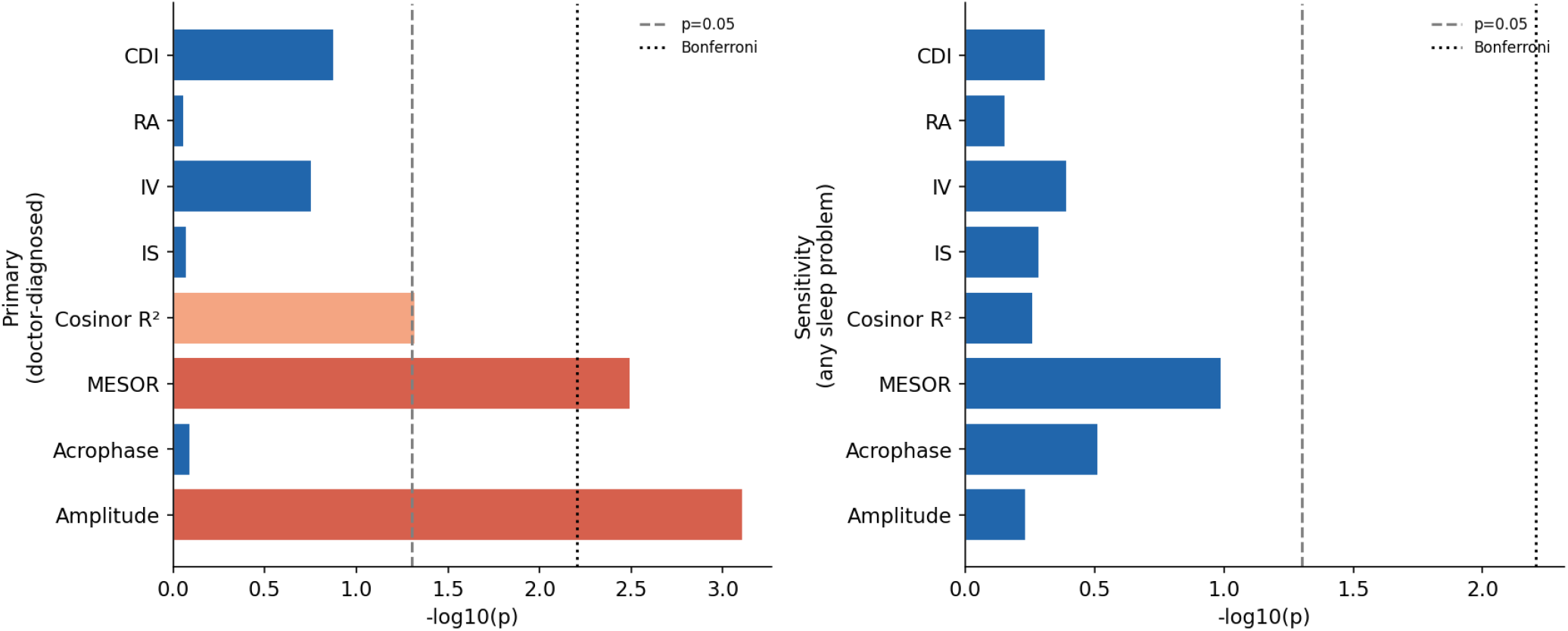
Survey-corrected Wald F-test results (−log_10_ *p*) for the age × sleep disorder interaction. Left panel: primary analysis (doctor-diagnosed disorder). Right panel: sensitivity analysis (any sleep problem, including self-reported). Dashed vertical line, nominal *p* = 0.05 threshold; dotted line, Bonferroni-corrected threshold (*p* = 0.006). Dark red bars are Bonferroni significant; light red bars are nominally significant; blue bars are non-significant.

### 3.4 CDI Associations with Cardiometabolic Outcomes

Table 3 and Figure 4 present associations between CDI and cardiometabolic outcomes adjusted for age, sex, and BMI. Each 1-SD increase in CDI was associated with higher BMI (*β* = 1.33 kg/m^2^, 95% CI [0.95, 1.70], *p* < 0.001), higher HbA1c (*β* = 0.089%, 95% CI [0.037, 0.141], *p* = 0.004), greater diabetes prevalence (*β* = 3.8 percentage points, 95% CI [2.2, 5.3], *p* < 0.001), and worse PHQ-9 depressive symptoms (*β* = 0.43 points, 95% CI [0.10, 0.76], *p* = 0.020).

**Table 3:** Associations between Circadian Disruption Index and cardiometabolic outcomes (survey-corrected).

| Outcome | $\beta$ | SE | 95% CI | $p$ | N |
| --- | --- | --- | --- | --- | --- |
| BMI (kg/m<sup>2</sup>) | 1.328 | 0.192 | [0.953, 1.704] | <0.001 | 4,386 |
| HbA1c (%) | 0.089 | 0.027 | [0.037, 0.141] | 0.004 | 4,213 |
| Diabetes (proportion) | 0.038 | 0.008 | [0.022, 0.053] | <0.001 | 4,385 |
| PHQ-9 score | 0.429 | 0.167 | [0.102, 0.757] | 0.020 | 3,821 |

$\beta$ coefficients represent change in each outcome per 1 SD increase in CDI, adjusted for age, sex, and BMI (BMI model excludes BMI as covariate). Standard errors and 95% CIs estimated via Taylor series linearization. PHQ-9, Patient Health Questionnaire-9.

**Figure 4:**
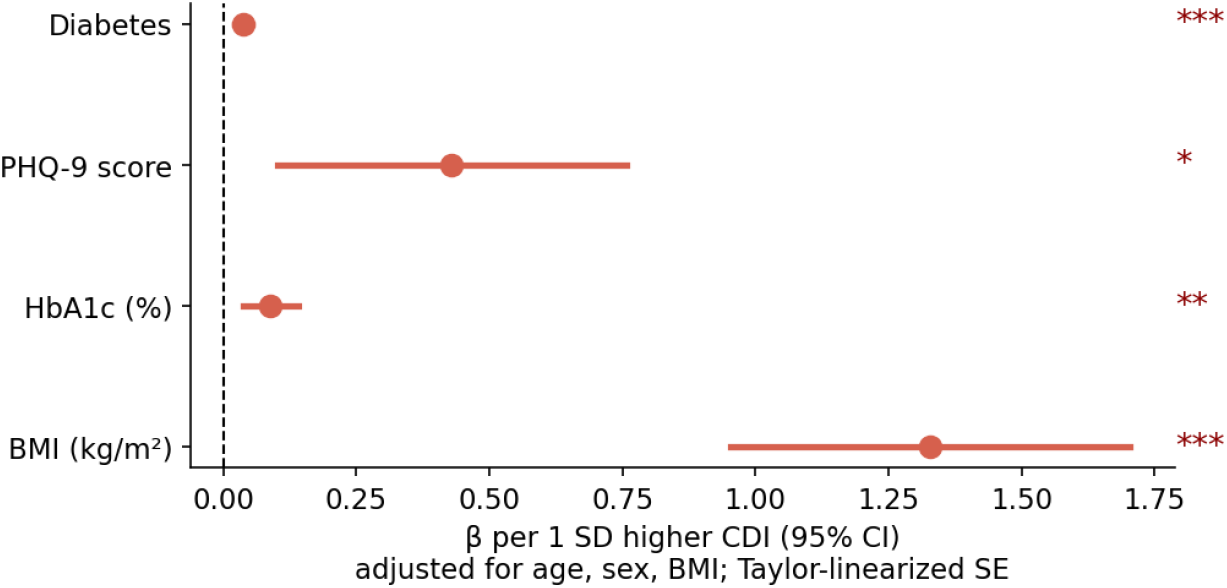
Forest plot of CDI associations with significant cardiometabolic and mental health outcomes (*p* < 0.05, survey-corrected). Points represent *β* coefficients (change per 1 SD CDI); horizontal lines represent 95% confidence intervals. Stars denote significance level (* *p* < 0.05, ** *p* < 0.01, *** *p* < 0.001). All models adjusted for age, sex, and BMI using Taylor-linearized survey variance.

### 3.5 Sensitivity Analysis

When the composite “any sleep problem” exposure (*N* = 1,087, 24.8%) was substituted for the primary exposure, no significant age-by-sleep-disorder interactions were observed for any circadian metric (all *p >* 0.05; Figure 3, right panel). This suggests that the primary findings are specific to individuals with a clinically diagnosed sleep disorder rather than those with self-reported sleep complaints more broadly.

## 4 Discussion

In this nationally representative study of 4,386 US participants, we found that doctor-diagnosed sleep disorder significantly modifies the age-related trajectory of circadian rest-activity amplitude and mean activity level (MESOR), with both interactions surviving Bonferroni correction. The CDI was robustly associated with four cardiometabolic and mental health outcomes after survey-corrected variance estimation. These findings advance understanding of the relationship between clinical sleep pathology and circadian aging in the general US population.

The significant age-by-sleep-disorder interactions for amplitude and MESOR indicate that the pattern by which these indices change across the lifespan differs by sleep disorder status. These differences are likely attributable to multiple converging mechanisms. Obstructive sleep apnea, the most prevalent clinically diagnosed sleep disorder in the general US adult population, produces intermittent hypoxia that disrupts peripheral circadian clock gene expression, activates systemic inflammation, and perturbs autonomic regulation of the rest-activity cycle [21–23, 25]. Chronic insomnia is characterized by sustained cortical and autonomic hyperarousal that fragments rest-activity rhythms even when total sleep duration is preserved [24, 48]. The observation that these effects manifest specifically on amplitude and MESOR, rather than on acrophase or fragmentation metrics, is consistent with evidence that the SCN pacemaker amplitude, rather than its period or phase, is the component most vulnerable to age-related degradation [2, 6, 10, 11].

The CDI associations with HbA1c, diabetes, PHQ-9 scores, and BMI replicate and extend prior observational findings linking circadian disruption to metabolic and psychiatric outcomes [14, 15, 48–51]. The magnitude of the HbA1c association (0.089% per 1 SD CDI) is clinically relevant relative to the diagnostic thresholds for prediabetes (5.7%) and diabetes (6.5%). Experimental evidence from controlled circadian misalignment protocols demonstrates that disruption of circadian timing per se, rather than total sleep restriction alone, drives acute elevations in insulin resistance and inflammatory markers [39–42], providing a plausible biological pathway. The PHQ-9 association is consistent with experimental evidence that circadian disruption directly impairs mood regulation and that circadian rhythm disturbances are a core feature of mood disorders [48, 52–54].

The failure of the sensitivity analysis to replicate the primary interaction findings when using the broader “any sleep problem” composite is informative. Doctor-diagnosed sleep disorders likely represent a more severe, biologically homogeneous, and chronically maintained phenotype than self-reported sleep complaints, which span a wide spectrum from transient insomnia to subclinical sleep dissatisfaction [17, 20]. The diagnostic specificity of the primary findings suggests that the threshold for circadian trajectory modification is a clinically significant, physician-confirmed condition rather than any sleep difficulty. This aligns with findings from other domains in which the health consequences of sleep problems are more pronounced in those with formal diagnoses versus self-report alone [19, 55, 56]. Future studies with disorder-specific diagnoses (apnea, insomnia, restless legs syndrome, circadian rhythm disorders) would help delineate which sleep disorder subtypes drive the observed interaction.

Several limitations warrant consideration. The crosssectional design precludes causal inference: sleep disorders and circadian disruption may be bidirectionally reinforcing, with disrupted circadian organization worsening sleep quality and vice versa [45–47]. The accelerometer measures movement rather than core circadian biological markers such as dim-light melatonin onset or core body temperature, and rest-activity rhythms represent a behavioral output of the circadian system rather than its molecular underpinning [3, 57, 58]. The analytic sample represents approximately 63% of eligible actigraphy participants after completeness restrictions, which could introduce selection bias if protocol completion is related to sleep disorder status or circadian function. The analysis is limited to a single NHANES cycle (2011–2012), and statistical power for subgroup analyses is constrained by the sample size of the diagnosed sleep disorder group (*N* = 360). The sleep disorder exposure (SLQ060) was administered only to participants aged 16 and older in NHANES 2011–2012; children aged 6–15 were therefore classified as not having a doctor-diagnosed disorder by default, potentially biasing interaction estimates in the younger age stratum toward the null. Finally, the selfreported nature of the sleep disorder variable may misclassify some participants, likely attenuating true effect estimates toward the null.

## 5 Conclusions

Doctor-diagnosed sleep disorders are associated with significantly different age-related trajectories of circadian activity amplitude and mean activity level in a large, nationally representative US population sample. The CDI is independently associated with higher BMI, elevated HbA1c, greater diabetes prevalence, and worse depressive symptoms. These findings suggest that clinically significant sleep pathology alters the normal circadian aging process, with measurable downstream consequences for metabolic and mental health. Longitudinal studies with repeated actigraphy assessments, disorder-specific sleep diagnoses, and circadian biomarkers are needed to clarify the direction and mechanism of these associations and to identify windows for intervention.

## Data Availability

The data analyzed in this study are publicly available from the National Center for Health Statistics. NHANES 2011-2012 accelerometry and questionnaire data can be downloaded at no cost from https://wwwn.cdc.gov/nchs/nhanes/continuousnhanes/default.aspx?BeginYear=2011. Analysis code is available from the corresponding author upon reasonable request.

## Author Contributions

L.Y.: conceptualization, data curation, formal analysis, methodology, software, visualization, writing (original draft). C.L.: investigation, validation, writing (review and editing). A.W.: investigation, validation, writing (review and editing). All authors have read and agreed to the published version of the manuscript.

## Institutional Review Board Statement

The NHANES protocol was approved by the National Center for Health Statistics Research Ethics Review Board. Ethical review and approval were waived for this study as it constitutes a secondary analysis of publicly available, de-identified data.

## Informed Consent Statement

Informed consent was obtained from all subjects involved in the original NHANES data collection. Not applicable for this secondary analysis.

## Data Availability Statement

NHANES 2011–2012 data are publicly available from the US Centers for Disease Control and Prevention at https://www.cdc.gov/nchs/nhanes/ (accessed on 1 January 2026). All analysis code is available from the corresponding author upon reasonable request.

## Acknowledgments

The authors thank the National Center for Health Statistics and all NHANES 2011–2012 participants for making this research possible.

